# Childhood traumatic experiences and sleepwalking events in the lower GDP counties in Hungary

**DOI:** 10.1101/2024.03.14.24304295

**Authors:** Vivian M. Correa

**Affiliations:** Mental Health Sciences ‘Doctoral School, Department of Preventive Health Science, Széchenyi University of Győr, Hungary

**Keywords:** Sleepwalking, Risk of injury, Childhood trauma, Hungarian population

## Abstract

**Purpose:** The aim of the study was to identify injurious behavior during sleepwalking events and childhood traumatic experiences.

**Study design:** Eight questions from a sleepwalking study (02/2023) in different counties in Hungary were analyzed to find a relationship between traumatic experiences in the life of the participants, such as physical, emotional, or sexual abuse in childhood, and injurious behaviors during sleepwalking.

**Methods:** We estimated an association between the Hungarian counties by GDP per capita and individuals that reported injurious sleepwalking events and childhood trauma by logistic regression.

**Results:** A total of 749 participants agreed to answer all questions, of which 357 were over 50 years old and 363 males. The results have shown that sleepwalking events were reported in patients with childhood trauma significantly more frequently in the lower GDP counties in Hungary (OR:5.8, 1.21-27.99, P:0.02) even without family predisposition.

**Conclusion:** The results suggest that a childhood traumatic experience could be a ssociated with sleepwalking activities. Thus, family history and traumatic experiences can increase the probability of injurious behavior during sleepwalking in Hungary.

## Introduction

Sleepwalking is a parasomnia that occurs during non-rapid eye movement (NREM) sleep, leading individuals to engage in complex behaviors while remaining asleep [1]. Some literature suggests a bidirectional relationship between sleep disturbances and childhood traumatic experiences [2]. The Diagnostic and Statistical Manual of Mental Disorders (DSM-5) categorizes traumatic events as “Trauma and Stress-Related Disorders” [3]. Include acute stress disorder, posttraumatic stress disorder (PTSD), and adjustment disorders [4]. Historically, insomnia and nightmares have been viewed as symptoms of PTSD disorder. Individuals who have experienced more severe trauma-related factors such as family loss, personal injury, etc., would exhibit more severe sleep disorders than those experiencing fewer trauma-related factors. Recent research has shown a significant association between childhood abuse and sleepwalking in the adult population [5]. Sleepwalking is most common in childhood and decreases with age with a prevalence ranging around 7% in certain parts of the world [6]. However, some factors such as socioeconomic status (race/ethnicity, media, and physical environment) influence not just in sleep but in overall health [7]. Thus, macro-, and micro-environmental factors can interfere with this aspect. Macro-environment refers to a condition in the country economy, such as the gross domestic product (GDP), inflation, and unsatisfactory political decisions (fiscal and monetary policy), which can act as a mediator of income inequality, and health-related quality of life [8]. Meanwhile, micro-environmental is related to a severe distress such as unemployment, family dissolution, lack of education, and/or drug abuse which can play a role as a risk factor for adults’ long-term sleep problems [9]. Traumatic experiences can activate the stress response system, impair emotion regulation, and fragment memory, which contributes to the onset or exacerbation of sleepwalking episodes. There is no childhood abuse versus somnambulism research conducted in Hungary in the literature. Here, we aim to assess the prevalence of sleepwalking and traumatic events in our population by social demographics levels.

## Methodology

This study investigated the sleepwalking prevalence in different Hungarian counties targeting both genders of the population from 19-year-old. We screened the ZRI Závecz Research database, Budapest, Hungary from February 28 to March 8, 2023, (n:1,000) for childhood traumatic experience associated with somnambulism. Participants were extracted anonymized, double-checked for Informed consent to be included in our investigation.

### Study participants

A total of 1,000 participants were voluntarily interviewed after their registration in a survey agency company. The survey agency collected the data based on a convenient sample of Hungarian counties, and all participants have given written informed consent.

### Data collection

The sleepwalking survey was performed by ZRI Závecz Research agency from February 28 to March 8, 2023, in Hungary. The questionnaire was elaborated by two experts with yes or no choices, and open-ended questions for the data collection (supplementary data - Table1). The open-ended questions intended to measure what the type of activities was performed in a sleepwalking event and childhood traumatic experiences reports. The interview was made in the local language.

### Statistical analysis

The data were analysed through Statistical Package for Social Sciences (SPSS). Percentages and chi-square test was utilized to represent statistical differences between the variables. Odds ratio at CIs 95% to find an association between the Hungarian counties by GDP per capita and individuals that reported sleepwalking events and childhood trauma.

### Exclusion criteria

Participants should have answered all the 8 questions. If one of the questions were preferred not to be answered, this participant was excluded.

### Limitations

The use of sleep aids or diagnosed sleep disorders, were not investigated. Consideration of other factors could establish a more robust and comprehensive understanding of the topic. However, the aim here was to calculate the probability of a patient to have injurious behaviour during sleepwalking events.

## Result

Of 749 screened, 8 were associated with traumatic experiences and sleepwalking. Three participants (37%) reported injurious activities during sleepwalking, and family history of known non-rapid eye movement parasomnias.

### Demographic data

Of the 1,000 Hungarians participants, 749 answered all the 8 questions, representing a response rate of 75%. The sample consisted of 181 (24%) from the capital Budapest and 568 (76%) from the countryside, with an approximately equal proportion of female individuals 396 (53%) and age raging between 19 and 80 years (median = 47y). Hungary has 19 counties, and from that 7 counties were excluded by the exclusion criteria. The remaining 12 Hungarian counties were divided according to GDP per capita in group 1 (GDP higher than 25k), and group 2 (GDP less than 25k), as shown on supplementary data Table 2 and 3.

Twenty of the 749 participants (2.7%) reported sleepwalking in the population. Three of the 8 (37%) by the respective lower GDP group reported sleepwalking events after having a childhood traumatic experience, but the difference was not statistically significant (X^2^ 2.8, df1, p:0.09). Forty-five of the 62 individuals who had experienced trauma (72%) were in the GDP higher than 25 k group, however, five of 17 childhood traumatic experience reported to start sleepwalking in the GDP less than 25 k group. The logistic regression results showed an association among sleepwalking events after a childhood traumatic experience, OR:5.8, 1.21-27.99, P:0.02, in the lower GDP counties group, when using trauma as a reference group (Table 1).

**TABLE 1.** Frequency of injuries and family disposition in sleepwalking participants, showing the binary logistic regression association between sleepwalking events, and childhood trauma by counties groups (N=749)

| Counties groups n (%) | Sleepwalking | Both | Trauma | X2, P-value | OR, P-value |
| --- | --- | --- | --- | --- | --- |
| **Group 1 (higher GDP)** | 12 (60) | 3 (37) | 45 (72) | 2.8, 0.09 | 5.0, 0.10 |
| **Group 2 (lower GDP)** | 8 (40) | 5 (62) | 17 (27) | 5.0, 0.10 | 5.8, 0.02* |
| **Total population n (%)** | 20 (2.7) | 8 (40) | 62 (8.3) |  |  |
| Injury in the total of SW | 6 (30) | 6 (75) |  |  |  |
| Family predisposition | 6 (30) | 3 (37) |  |  |  |

\*Statistically significant

## Discussion

The prevalence of sleepwalking in Hungary is not well documented, but studies suggest that they are common and may affect a significant proportion of the population. In our results the sleepwalking prevalence in Hungary was 2,7%, which is in accordance with the worldwide data ranging from 2 to 7%. However, here we are discussing that sleepwalking can be considered a protective mechanism more likely to happen when the patient has a history of psychological trauma, as we see in the literature [10].

The results have shown forty percent of the participants reported sleepwalking and traumatic experiences, such as being aggressively beaten by a drunk father, or lost someone in the family. Losing a family member can be emotionally devastating, resulting in profound grief, and a range of psychological reactions. These results are in line with a study conducted by Hartman D. et al., where a minority of adult patients had a history of major psychological trauma and were sleepwalkers [10].

The high probability of injurious activities during sleepwalking after childhood traumatic experiences was 5.8-fold associated with the low GDP per capita group. Even though 37% of the participants reported the same in the higher GDP group, we must take into consideration this is a delicate topic and it cannot be comfortable for some participants to answer. However, we made sure participants understood all the instructions and we provided information about the study in the local language.

The strength of this research was to present data gathered with the literature analyses including variables such as childhood trauma and emotional distress which can play a role in the injurious behaviors during sleepwalking events. In this context, treatments addressing both sleep and emotional problems are essential. A good intervention strategy to improve participants’ sleep quality in future interventions should consider the link between childhood trauma and quality of life. A simple list of recommendations for better sleep would not be enough, it would be necessary to provide adequate knowledge and resources. Here, the high prevalence of injuries during sleepwalking reported after childhood trauma was remarkable.

## Conclusion

Childhood traumatic experiences can have long-lasting effects on mental and physical health and may be linked to sleep disorders such as sleepwalking in adulthood. Family history is particularly important in the sleepwalking prevalence in adults, specially in the prevention of injurious behaviors during sleepwalking.

## Data Availability

The data used to support the findings of this study were provided by the institution. Access to these data should be requested from the author.

## Acknowledgment

*Tempus Public Foundation, Stipendium Hungaricum program*.

## Annex 1 supplementary data

**TABLE 1.** Questionnaire on a sample of 1,000 people representing the country’s adult population, using a personal data collection method.

| Questions | Type |
| --- | --- |
| 1. What is your age? | open-ended |
| 2. What is your biological sex? | female - male |
| 3. In which Hungarian county do you live? | open-ended |
| 4. Have you ever started walking or doing some activity while you were sleeping? | yes or no |
| 5. What did you usually do during the sleepwalking episode? | open-ended |
| 6. Have you had any disturbing, or injurious behavior while you were sleeping? | open-ended |
| 7. Has anyone in your family been diagnosed with a sleep disorder? | yes or no |
| 8. Have you suffered any serious mental abuse or harm as a child or younger? | open-ended |

**TABLE 2.** GPS coordinates in the Hungarian counties by GDP per capita in USD in 2022 according to data by the OECD *.

| **Group 1 – countryside plus capital** | **GDP higher than 25k** | **GPS coordinates** | **N** |
| --- | --- | --- | --- |
| Budapest - Capital | 64,298 | 47°29'33"N 19°03'05"E | 181 |
| Győr-Moson-Sopron | 38,449 | 47°40'0.01" N 17°15'0.00" E | 49 |
| Komárom-Esztergom | 32,158 | 47°34'59.99" N 18°19'59.99" E | 28 |
| Fejér | 32,158 | 47°10'0.01" N 18°34'59.99" E | 43 |
| Vas | 28,667 | 47°10'0.01" N 16°45'0.00" E | 27 |
| Pest | 25,023 | 47°25'0.01" N 19°19'59.99" E | 119 |
| Bács-Kiskun | 25,718 | 46°30'0.00" N 19°25'0.01" E | 51 |
| Total |  |  | 498 |
| **Group 2 - counties** | **GDP less than 25k** |  |  |
| Tolna | 23,778 | 46°30'0.00" N 18°34'59.99" E | 21 |
| Heves | 23,566 | 47°49'59.99" N 20°15'0.00" E | 28 |
| Borsod-Abaúj-Zemplén | 23,249 | 48°15'0.00" N 21°00'0.00" E | 66 |
| Csongrád-Csanád | 23,218 | 46°25'0.01" N 20°15'0.00" E | 45 |
| Hajdú-Bihar | 22,678 | 47°25'0.01" N 21°30'0.00" E | 54 |
| Békés | 18,938 | 46°45'0.00" N 21°00'0.00" E | 37 |
| Total |  |  | 251 |

\* https://www.oecd.org/regional/HUN-RCG2022.pdf

**TABLE 3.** Hungarian counties used in the research by region and GDP per capita in 2022 according to the OECD data.

|  | County | Order of counties on the basis of GPD per capita in 2022 |
| --- | --- | --- |
| **Budapest - Capital region\*** |  | 1 st |
| **Central Hungary** | Pest - County region* | 6 nd |
| **Central Transdanubia** | Fejér* | 3 rd |
|  | Komárom-Esztergom* | 4 th |
|  | Veszprém | 9 th |
| **Western Transdanubia** | Győr-Moson-Sopron* | 2 th |
|  | Vas* | 5 th |
|  | Zala | 8 th |
| **Southern Transdanubia** | Baranya | 16 th |
|  | Somogy | 17 th |
|  | Tolna* | 11 st |
| **Northern Hungary** | Borsod-Abaúj-Zemplén * | 13 rd |
|  | Heves* | 12 nd |
|  | Nógrád | 20 th |
| **Northern Great Plain** | Hajdú-Bihar * | 14 th |
|  | Jász-Nagykun-Szolnok | 15 th |
|  | Szabolcs-Szatmár-Bereg | 19 th |
| **Southern Great Plain** | Bács-Kiskun* | 7 th |
|  | Békés* | 18 th |
|  | Csongrád-Csanád | 10 th |

\*Counties used in the database

